# Ventilation rate assessment by carbon dioxide levels in dental treatment rooms

**DOI:** 10.1101/2021.02.04.21251153

**Authors:** Qirong Huang, Tamer Marzouk, Razvan Cirligeanu, Hans Malmstrom, Eli Eliav, Yan-Fang Ren

## Abstract

**Objectives:** The purpose of the present study was to monitor and evaluate CO_2_ levels in dental operatories using a consumer-grade CO_2_ sensor and determine the utility and accuracy of various methods using CO_2_ levels to assess ventilation rate in dental clinics. We aim to find a practical tool for dental practitioners to conveniently and accurately monitor CO_2_ levels and assess the ventilation rates in their office in order to devise a pragmatic and effective strategy for ventilation improvement in their work environment.

**Methods:** Mechanical ventilation rate in air change per hour (ACH_VENT_) of 10 dental operatories was first measured with an air velocity sensor and air flow balancing hood. CO_2_ levels were measured in these rooms to analyze the effects of ventilation rate and number of persons in the room on CO_2_ accumulation. Ventilation rates were estimated using natural steady state CO_2_ levels during dental treatments and experimental CO_2_ concentration decays by dry ice or mixing baking soda and vinegar. We compared the differences and assessed the correlations between ACH_VENT_ and ventilation rates estimated by steady states CO_2_ model with low (0.3 L/min, ACH_SS30_) or high (0.46 L/min, ACH_SS46_) CO_2_ generation rates, by CO_2_ decay constants using dry ice (ACH_DI_) or baking soda (ACH_BV_), and by time needed to remove 63% of excess CO_2_ generated by dry ice (ACH_DI63%_) or baking soda (ACH_BV63%_).

**Results:** ACH_VENT_ varied from 3.9 to 35.0 with a mean of 13.2 (±10.6) in the 10 dental operatories. CO_2_ accumulation occurred in rooms with low ventilation (ACH_VENT_≤6) and more persons (n>3) but not in those with higher ventilation and less persons. ACH_SS30_ and ACH_SS46_ correlated well with ACH_VENT_ (r=0.83, p=0.003), but ACH_SS30_ was more accurate for rooms with low ACH_VENT_. Ventilation rates could be reliably estimated using CO_2_ released from dry ice or baking soda. ACH_VENT_ was highly correlated with ACH_DI_ (r=0.99), ACH_BV_(r=0.98), ACH_DI63%_(r=0.98), and ACH_BV63%_ (r=0.98). There were no statistically significant differences between ACH_VENT_ and ACH_DI63%_ or ACH_BV63%_.

**Conclusions:** Dental operatories with low ventilation rates and overcrowding facilitate CO_2_ accumulations. Ventilation rates could be reliably calculated by observing the changes in CO_2_ levels after a simple mixing of household baking soda and vinegar in dental settings. Time needed to remove 63% of excess CO_2_ generated by baking soda could be used to accurately assess the ventilation rates using a consumer-grade CO_2_ sensor and a basic calculator.

## Introduction

Risks of disease transmission in healthcare settings during infectious disease pandemics have consistently challenged dental care professionals (DCPs) in their efforts to maintain a safe environment for their staffs and patients. DCPs have gained tremendous experiences in infection control from the ongoing HIV/AIDS pandemic by implementing universal or standard precautions against contact and droplet transmissions, but we are less confident in dealing with an infectious respiratory disease that may be transmitted through aerosol particles emitted by patients who have no overt symptoms. With mounting evidence that COVID-19 is transmissible through aerosols in an indoor environment [1, 2], additional preventive measures beyond the standard care using personal protective equipment (PPE) are essential to minimize risks and alleviate anxieties experienced by staff and patients due to uncertainties associated with a novel infectious respiratory disease pandemic.

Engineering controls through mechanical ventilation are important mechanisms to reduce the risks of airborne disease transmission in an indoor environment such as the dental offices.

Though CDC recommends improving ventilation and air filtration in its guidance for dental settings during the COVID-19 pandemic [3], few information is available on how to assess the ventilation condition and what measures to take to achieve more effective engineering control of disease transmission in dental offices.

Carbon dioxide (CO_2_) level is an important indicator of ventilation in occupied indoor environments. CO_2_ is a byproduct of human metabolism and exists in high levels in exhaled air. Atmospheric CO_2_ level is at approximately 400 parts per million (ppm) in most outdoor environments, but CO_2_ in human exhaled air reaches on average 40,000 ppm in concentration [4]. CO_2_ levels is therefore often used as a proxy for indoor air quality as well as a risk marker for transmission of airborne diseases since it is inert, its indoor emission source (human) is known, and its measurement is inexpensive and accurate [5]). Accumulation of CO_2_ may occur in indoor spaces with poor ventilation and overcrowding, and consequentially increase the risks of disease transmission, which is of particular significance under the current COVID-19 pandemic. High levels of CO_2_ in indoor environments have been associated with the transmission of infectious respiratory diseases such as tuberculosis, influenza and rhinovirus infections [6-8]. Indoor CO_2_ levels have therefore been widely used to model the risks of airborne infectious disease transmission [4, 9, 10], including that of coronavirus infection transmission in dental offices [11].

CO_2_ levels is directly associated with ventilation rate in an occupied space. A clinical space with good ventilation should have a CO_2_ level close to that of outside air at approximately 400 ppm in most areas [4, 12]. Higher indoor CO_2_ levels indicate poor ventilation, accumulation of exhaled air, and increase in the fraction of “rebreathed air” or “shared air” in the indoor environment, which is proven to be a risk factor for transmissions of respiratory infectious diseases [4, 7, 8, 10]. CO_2_ levels have been used to estimate ventilation rates in dental offices through mathematic modeling [13-15]. Godwin and colleagues reported that the ventilation rate was 1.12 air change per hour (ACH) in a typical dental clinic in the US [13], and Helmis and colleagues found that it was on average 5 ACH in a dental school clinic in Greece where the doors and windows were opened for cross ventilation [15]. Both studies used natural build-up of CO_2_ levels in the dental clinic to estimate the ventilation rate through mathematic models, but none of them actually verified the ventilation estimates using conventional methodologies such as a high-precision airflow sensor [16]. It is not known if these estimates were accurate as both methods require mathematical modeling based on several assumptions related to CO_2_ generation, build-up and dispersion over a relatively lengthy period of time, which may result in erroneous estimates if any of the assumed conditions are not met [5]. A simpler and more reliable method is needed if CO_2_ level is to be used by dental practitioners to assess the ventilation rate of their treatment rooms.

The purpose of the present study was two-folds: 1) to monitor and evaluate CO_2_ level and its associated factors in dental operatories using a consumer-grade CO_2_ sensor, and 2) to determine the utility and accuracy of various methods that use CO_2_ levels to assess ventilation rate in dental clinics. Our aim was to find a practical tool that will enable all dental practitioners to conveniently and accurately monitor CO_2_ levels and assess the ventilation rates in their treatment rooms in order to devise a pragmatic and effective strategy for ventilation improvement in their work environment.

## Methods

### Study settings

We conducted the CO_2_ concentration and ventilation rate assessments in 10 treatment rooms ranging from 667 to 1221 cubic feet (ft^3^) in sizes. Mechanical ventilations of the treatment rooms are provided by three separate air handlers that drew 30%, 50% and 50% outside air to the ventilation systems. The outside air percentage were later increased to 60% for all three air handlers as part of our institutional response to COVID-19.

### Determining room airflow and mechanical ventilation rates

The mechanical ventilation of the 10 selected dental treatment rooms was measured with an air velocity sensor integrated in an air flow balancing hood (ADM-850L Airdata Multimeter with CFM-850L FlowHood, Shortridge Instruments Inc., Scottsdale, AZ) as described elsewhere [17]. Briefly, the volumetric airflow rates of the dental treatment rooms were measured in cubic feet per minute (CFM, or ft^3^/min) at both the air supply inlets and air exhaust returns using the air flow balancing hood. The mechanical ventilation rates of each space in number of ACH was calculated based on supply and exhaust airflow rates for each room. The larger value between ACH from supply air and ACH from return air was used as the room’s mechanical ventilation rate (ACH_VENT_) [18].

### Assessing CO_2_ levels during dental treatment procedures

We measured CO_2_ levels in two dental treatment rooms when dental procedures were performed. The two rooms represented two extremes in ventilation rates, with one at the lowest at 3.9 ACH, and the other at the highest at 35 ACH. The number of persons in the rooms were recorded in real time when a person was entering and leaving the room. One of the two rooms were used for dental implant therapy and may often have more than one graduate students observing the procedures in addition to the treating dentist, the dental assistant and the patient in the room.

### 24-hour Continuous monitoring CO_2_ in dental treatment rooms

To further explore the dynamics of CO_2_ levels in dental treatment rooms throughout the work day and assess of accuracies of the steady state models of CO_2_ for ventilation assessments, we continuously measured the CO_2_ levels in the 10 selected dental treatment rooms for 24 hours and recorded the procedures performed and number of persons in the room.

### Assessing ventilate rate by natural CO_2_ level modeling in dental treatment rooms

We used the steady state model described by Batterman [5] to calculate the ACH of the treatment rooms and compared the outcomes with ACH_VENT_ determined by mechanical ventilation.

The steady state air change rate (ACH_SS_) is calculated as follows [5]:

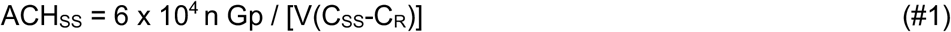

Where n = number of persons in the room; G_P_ = average CO_2_ generation rate; V = volume of the room in cubic meters (m^3^); C_SS_ = steady state indoor CO_2_ level in ppm; and C_R_ = CO_2_ level in outdoor air in ppm.

The CO_2_ generation rate G_P_ is affected by many factors and may vary by human activity, physical size, sex and race [19, 20]. G_P_ values of 0.46 L/min and 0.30 L/min have been used in previous studies to represent CO_2_ generation by moderately active adults [5], [13]). As the CO_2_ generation rates and activity levels by dental care providers and their patients are unknown and may not be constant, we decided to use both values to calculate the steady state air change rate ACH_SS_ and assess the correlations between ACH_SS_ and ACH_VENT_ at two G_P_ levels (0.46 and 0.30 L/min, respectively).

### Assessing ventilation rates by CO_2_ decays using dry ice

Ventilation rates were also measured in the 10 dental treatment rooms using the CO_2_ decay method [5]. Outside air CO_2_ level was first measured for 5 minutes near the air intake of the ventilation system outside the building before each experiment. To raise the peak CO_2_ levels inside the dental treatment rooms to approximately 2000 ppm, 250g of dry ice were placed in a water bath and left in the room for two minutes. A small oscillating fan was used to keep the CO_2_ well mixed in the room. CO_2_ level was then measured at one-minute interval using an Aranet4 CO_2_ sensor (range 0-9999ppm, accuracy ±50ppm, SAF Tehnika, Riga, Latvia) for up two hours. The ventilation rate by CO_2_ clearance using dry ice (ACH_DI_) were determined as described by Batterman [5] using the CO_2_ concentration decays:

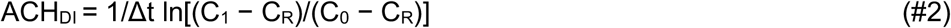

where Δt = period between measurements; C_0_ and C_1_ = CO_2_ levels measured at the beginning and the end of the decay period (ppm), and C_R_ = CO_2_ level in outdoor air (ppm). The consumer-grade Aranet4 CO_2_ sensor used in the current study was purchased at amazon.com in the US. It was recommended by the Federation of European Heating, Ventilation and Air Conditioning Associations (REHVA) for monitoring CO_2_ levels in schools during the COVID-19 pandemic [21], and was found to be comparable to a research-grade LI-COR CO_2_ sensor in accuracy and suitable for the time-response assessment in this study [22].

### Assessing ventilation rates by CO_2_ decays using baking soda

Considering that dental practitioners in private practices may not have ready access to dry ice, we developed a method to rapidly generate CO_2_ in dental treatment rooms using household baking soda (Arm& Hammer Pure baking soda, Church & Dwight Co., Inc., Ewing, NJ, USA) and vinegar (Heinz all-natural distilled white vinegar, Pittsburgh, PA, USA). Mixing baking soda (NaHCO_3_) with vinegar containing 5% acetic acid (CH_3_COOH) will generate CO_2_ as follows:

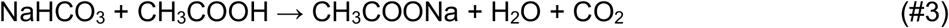

We tested peak CO_2_ values in the 10 treatment rooms using a weight (g):volume (ml) ratio of 1:15 based on the molar masses of the reagents. We aimed at a peak level range of 1500 to 2000 ppm in rooms with various mechanical ventilation rates and with doors closed. We determined that adding about 125g of baking soda (approximately 3/5 cup measure) to 1893 ml (a 64-oz bottle) of vinegar containing 5% acetic acid will elevate the CO_2_ level in a typical dental treatment room (10 x 11 ft in area, 8 ft in ceiling height, or 880 ft^3^ in volume) with a moderate ventilation rate (ACH_VENT_ = 4) to above 1500 ppm. For rooms that are significantly larger or having very high ventilation rates, one full cup measure (about 8 oz or 227g) of baking soda may be used with 3785 ml (a one-gallon jar) of vinegar containing 5% acetic acid.

Before the baking soda experiment, outdoor CO_2_ level was first measured for 5 minutes near the air intake of the ventilation system outside the building. To generate CO_2_ inside the dental treatment rooms, we first poured vinegar into a large container and added the baking soda powder. The mixture was vigorously shaken or stirred with a large spatula for two minutes and then removed from the room. A small oscillating fan was used to keep the CO_2_ well mixed in the room with doors closed. CO_2_ level was then measured at one-minute interval using the CO_2_ sensor as described above. The length of the CO_2_ measurement period is dependent on the ventilation rates of the room. For rooms with excellent ventilation, CO_2_ level decays rapidly and a measurement period of 30 minutes may be adequate. For rooms with very poor ventilation, CO_2_ measurements may need to continue for several hours or overnight to allow a sufficient level of CO_2_ clearance for calculating the ventilation rate using the baking soda and vinegar method (ACH_BV_) based on equation #2 described above.

### Estimating ventilation rate by time to 63% removal of excess CO_2_

Based on the commonly used formula for the rate of purging airborne contaminants, one complete air change will replace 63% of the airborne contaminants in the room with outdoor air [22-24]. Ventilation rate can therefore be simply calculated using the time needed to reach 63% reduction of excess CO_2_ from its peak level:

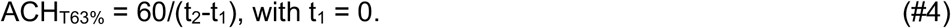

where t_1_ = initial timepoint in minutes with indoor CO_2_ at peak level, and t_2_ = timepoint in minutes when excess CO_2_ is reduced by 63%. Indoor CO_2_ at peak level (C_S_) is the sum of outdoor CO_2_ (C_R_) and excess CO_2_ (C_E_) generated by dry ice or baking soda. As CO_2_ measurement starts at peak level in this experiment, t_1_ is therefore always 0. Time needed to remove 63% C_E,_ or t_2,_ is the timepoint when indoor CO_2_ level is at C_63%E_ = C_S_ −63% C_E_, where C_E_ = C_S_ -C_R_.

### Statistical analysis

To understand factors associated with CO_2_ levels in dental treatment rooms, we performed multiple regression analysis using CO_2_ levels as dependent variable and number of persons in the room, ventilation rate, room sizem and outdoor CO_2_ level as independent variables. We analyzed the dynamics of CO_2_ levels during dental treatment procedures using descriptive analysis and compared the differences in steady state CO_2_ levels between rooms with poor and good ventilations using t-tests. ACH_VENT_ was compared with ACH calculated with different methods based on CO_2_ levels to assess the correlation (Pearson’s r) between the two methods of ventilation rate assessments. As the primary goal of the present study was to determine the validity of pragmatic methodologies that could be used by dental practitioners who do not have access to sophisticated instruments to assess the ventilation rate of their clinic spaces, we focused the analysis on CO_2_ generation using baking soda and vinegar and ventilation rate calculation using time needed to achieve 63% reduction of peak CO_2_ levels (ACH_BV63%_).

## Results

### Mechanical ventilation rate of the dental treatment rooms

The volumetric sizes, airflow rates and mechanical ventilation rates of dental treatment rooms are presented in Table 1. The 10 operatories are on average 882 ft^3^ in volume (range 667 to 1221 ft^3^). ACH_VENT_ varied from 3.9 to 35.0 with a mean of 13.2 (±10.6). A majority (7/10) of the treatment rooms have greater supply than exhaust air flow rates with positive differentials between ACH_s_ and ACH_E_ at two or greater; and one room have significantly greater exhaust than supply airflow rates with a highly negative differential between ACH_s_ and ACH_E_. A majority of the treatment rooms (7/10) have ACH_VENT_ at or greater than 6, and 4 of the 10 rooms had ACH_VENT_ greater than 10.

**Table 1:** Volumetric sizes and mechanical ventilation rates of dental operatories.

| Rm # | Volume<br>ft<sup>3</sup> | SAF<br>ft<sup>3</sup>/min | EAF<br>ft<sup>3</sup>/min | ACH<sub>s</sub> | ACH<sub>E</sub> | ACH<sub>VENT</sub> | Floor |
| --- | --- | --- | --- | --- | --- | --- | --- |
| 002 | 815 | 82 | 27 | 6.0 | 2.0 | 6.0 | 0 |
| 003 | 787 | 69 | 27 | 5.3 | 2.1 | 5.3 | 0 |
| 008 | 1221 | 149 | 103 | 7.3 | 5.1 | 7.3 | 2 |
| 012 | 1015 | 152 | 64 | 9.0 | 3.8 | 9.0 | 2 |
| 019 | 686 | 59 | 400 | 5.2 | 35.0 | 35.0 | 1 |
| 021 | 861 | 75 | 51 | 5.3 | 3.6 | 5.3 | 1 |
| 022 | 833 | 55 | 46 | 3.9 | 3.3 | 3.9 | 1 |
| 031 | 962 | 337 | 210 | 21.0 | 13.1 | 21.0 | 2 |
| 032 | 667 | 289 | 220 | 26.0 | 19.8 | 26.0 | 2 |
| 033 | 970 | 211 | 220 | 13.1 | 13.6 | 13.6 | 2 |
| **Mean** | 882 | 148 | 137 | 10.2 | 10.1 | 13.2 | - |
| **SD** | 166 | 101 | 122 | 7.6 | 10.6 | 10.6 | - |

SAF: supply airflow rate in cubic feet per minute. EAF: exhaust airflow rate in cubic feet per minute. ACH<sub>s</sub>: air change per hour based on supply airflow rate. ACH<sub>E</sub>: air change per hour based on exhaust airflow rate. ACH<sub>VENT</sub>: air change per hour based on mechanical ventilation. OA%: outside air percentage drawn by the air handlers.

### CO_2_ levels during dental treatment procedures

As shown in Figure 1, CO_2_ levels were significantly higher in the room with low ventilation rate (ACH = 3.9) and reached a peak of nearly 1600 ppm when 6 persons were in the room. The increased number of persons were related to teaching activities involving dental implant surgery where additional graduate students were allowed to observe the procedures. Comparing the two rooms with the same number of persons in the room for the same restorative procedures, CO_2_ levels reached 1100 ppm at the peak in the room with 3.9 ACH but stayed below 700 ppm in the room with 35 ACH (p<0.0001). CO_2_ accumulation appeared to be associated with crowding and low ventilation rate.

**Figure 1:**
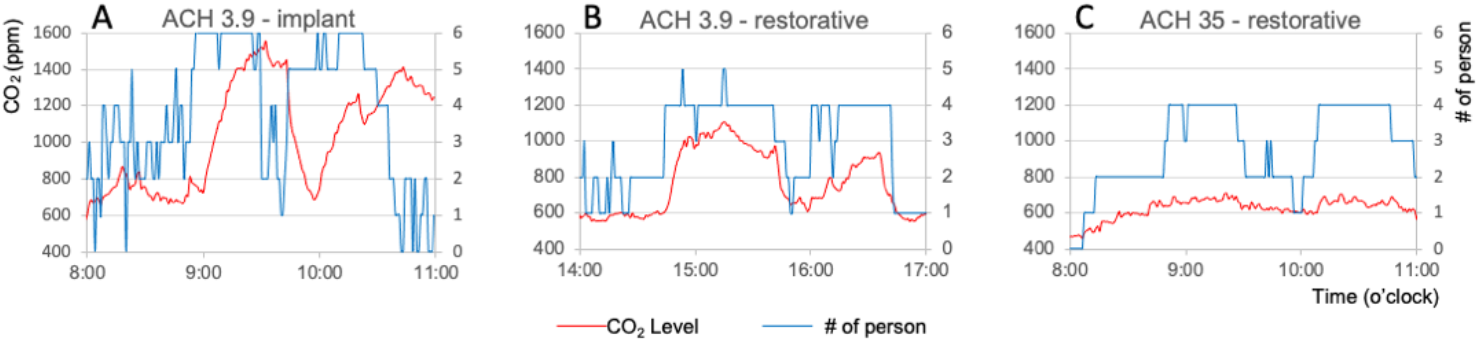
CO_2_ levels during dental treatment procedures in operatories with low and high mechanical ventilation. A: Significant CO_2_ accumulation occurred in a room with low ventilation (ACH = 3.9) and multiple persons in the room during clinical teaching activities for dental implant surgery. E, F: CO_2_ level is associated with ventilation rate in rooms with the same number of persons. Peak CO_2_ level reached 1100 ppm in the room with 3.9 ACH (B) but stayed under 700 ppm in the rooms with 35 ACH (C).

### Continuous CO_2_ monitoring in dental treatment rooms

We continuously monitored the CO_2_ levels for 24 hours in 10 treatment rooms with various ventilation rates in different departments. The working hours were from 8:00am to 5:00pm with lunch breaks from 12:30pm to 1:30pm for some and 1:00pm to 2:00pm for other departments. The dental procedures included prophylaxis, extractions, restoratives, endodontics, dental implant surgery, periodontal surgery and exams. Number of persons in the rooms varied from 2 to 6, with more people in the room during dental implant surgeries. The CO_2_ levels in early morning (5:00 - 7:00am) were at an level of 421±10 ppm, similar to the outdoor levels (413±15 ppm) (Table 2, Figure 2). The steady state CO_2_ level (C_SS_) during dental procedures, which was the mean concentrations of CO_2_ at the plateau level when the number of persons in the room stays unchanged for at least 5 min, ranged from 543 ppm to 1,374 ppm (786±207 ppm) in the 10 dental treatment rooms. Multiple regression analysis showed that steady state CO_2_ levels in dental treatment rooms were statistically significantly correlated to number of persons in the room (β=90.2, p=0.006), mechanical ventilation rate (β=11.0, p=0.001) and the volumetric size of the room (β=-0.50, p=0.049), but not to outside air CO_2_ levels (β=4.15, p=0.160). These findings confirmed that more people in smaller room with low ventilation rate facilitate CO_2_ accumulation in dental treatment rooms.

**Table 2:** Steady state CO_2_ levels during treatment procedures and ventilation rate estimates based on low (0.3 L/min) and high (0.46 L/min) assumptions of human CO_2_ generation rates.

| Rm # | Procedure | n<br>persons | C<sub>ss</sub><br>ppm | C<sub>R</sub><br>Ppm | ACH<sub>ss30</sub> | ACH<sub>ss46</sub> |
| --- | --- | --- | --- | --- | --- | --- |
| 002 | Exam | 4 | 1014 | 410 | 5.2 | 7.9 |
|  | Extraction | 4 | 978 | 410 | 5.5 | 8.4 |
| 003 | Extraction | 4 | 960 | 410 | 5.9 | 9.0 |
|  | Extraction | 4 | 923 | 410 | 6.3 | 9.7 |
| 008 | Hygiene | 2 | 673 | 435 | 4.4 | 6.7 |
|  | Hygiene | 2 | 629 | 435 | 5.4 | 8.2 |
| 012 | Hygiene | 2 | 649 | 434 | 5.8 | 8.9 |
|  | Hygiene | 2 | 632 | 434 | 6.3 | 9.7 |
| 019 | Extraction | 4 | 823 | 403 | 8.8 | 13.5 |
|  | Restorative | 3 | 616 | 403 | 13.1 | 20.1 |
| 021 | Implant | 4 | 905 | 410 | 6.0 | 9.1 |
|  | Endo | 4 | 926 | 410 | 5.7 | 8.8 |
| 022 | Implant | 6 | 1269 | 428 | 5.4 | 8.3 |
|  | Implant | 5 | 1089 | 428 | 5.8 | 8.8 |
| 031 | Hygiene | 2 | 544 | 405 | 9.5 | 14.6 |
|  | Surgery | 4 | 611 | 405 | 12.9 | 19.7 |
| 032 | Surgery | 3 | 584 | 404 | 15.8 | 24.2 |
|  | Exam | 4 | 633 | 404 | 16.7 | 25.5 |
| 033 | Exam | 3 | 595 | 392 | 9.7 | 14.8 |
|  | Surgery | 4 | 662 | 392 | 10.6 | 16.2 |
| **Mean** | - | 3.5 | 785.8 | 413.1 | 8.2 | 12.6 |
| **SD** | - | 1.1 | 206.8 | 14.0 | 3.8 | 5.8 |

Based on ventilation rate $ACH = 6 \times 10^4 n G_P / [V(C_{ss}-C_R)]$ ; C<sub>ss</sub>: steady state CO<sub>2</sub> level; C<sub>R</sub>: outdoor CO<sub>2</sub> level; ACH<sub>ss30</sub>: ventilation estimate based on CO<sub>2</sub> generation rate G<sub>P</sub>=0.3L/min per person; ACH<sub>ss46</sub>: ventilation estimate based on CO<sub>2</sub> generation rate G<sub>P</sub>=0.46L/min per person;

**Figure 2:**
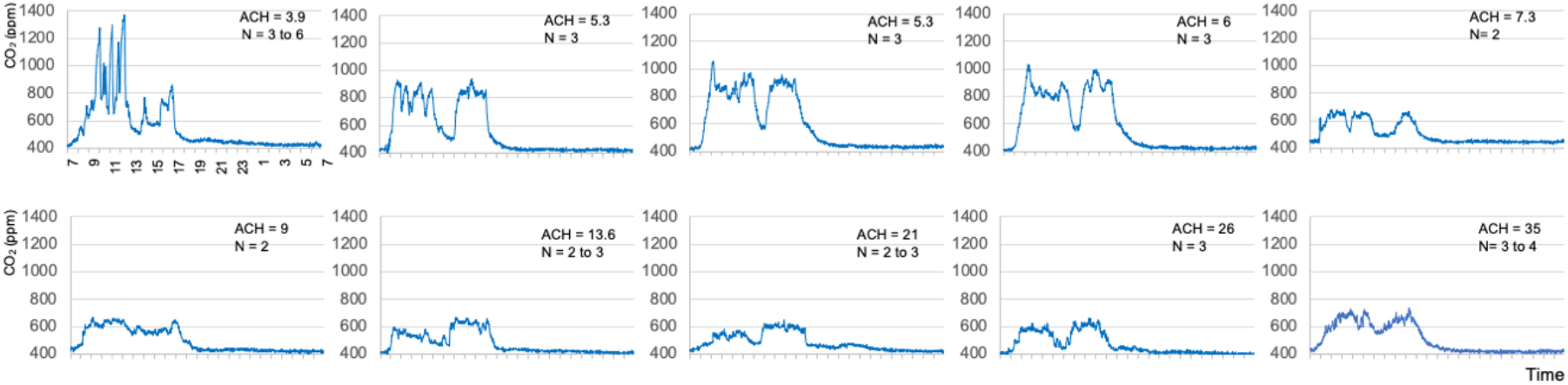
24-hour continuous measurements of CO_2_ levels in 10 dental treatment rooms with various ventilation rates. CO_2_ accumulation occurred in rooms with lower ventilation rates (ACH ≤ 6). CO_2_ levels stayed under 800 ppm in rooms with higher ventilation rate and lower number of persons. CO_2_ level in nonworking hours is close to that of outdoor at 400 ppm in all the rooms.

As shown in Figure 2, CO_2_ levels in rooms with ACH higher than 6 rarely reach 800 ppm. In rooms with ACH lower than 6, however, the CO_2_ levels were consistently greater than 800 ppm and approached 1,400 ppm in a room with ACH 3.9 when number of persons in the room was as high as 6 during clinical teaching activities related to dental implant procedures.

### Ventilation rates by steady state CO_2_ level modeling in dental treatment rooms

Based on steady state CO_2_ level (C_SS_) and outdoor CO_2_ level (C_R_) presented in Table 2, ventilation rates with CO_2_ generation at 0.30 L/min (ACH_SS30_) and 0.46 L/Min (ACH_SS46_) in the clinical space were calculated using equation #1 for two dental treatment procedures in each room. As expected, ACH_SS30_ values (Mean 8.3, SD 3.7) were significantly lower than ACH_SS46_ (Mean 12.6, SD 5.7) (mean difference = −4.5, paired t=-6.97, p<0.0001). Both ACH_SS30_ and ACH_SS46_ were similarly well correlated with ACH_VENT_ (r=0.83, p=0.003). ACH_SS30_ approximated closely to the mechanical ventilation rates in rooms with ACH_VENT_ ≤ 6 (mean difference=-0.6, paired t=-1.24, p=0.304), but significantly underestimated those in rooms with ACH_VENT_ > 6 (mean difference=-8.7, paired t=-2.59, p=0.049). The opposite is true for ACH_SS46_, it significantly overestimated the ventilation rates in rooms with ACH_VENT_ ≤ 6 (mean difference=3.7, paired t=6.78, p=0.007), but approximated closer to those in rooms with ACH_VENT_ > 6 (mean difference=-3.5, paired t=-1.14, p=0.307) (Table 3).

**Table 3:** Comparisons between mechanical ventilation rates and ventilation rates estimated from natural CO_2_ levels and CO_2_ released by dry ice and baking soda.

| Rm # | ACH<sub>VENT</sub> | ACH<sub>SS30</sub> | ACH<sub>SS46</sub> | ACH<sub>DI</sub> | ACH<sub>BV</sub> | ACH<sub>DI63</sub> | ACH<sub>BV63</sub> |
| --- | --- | --- | --- | --- | --- | --- | --- |
| 002 | 6.0 | 5.4 | 8.2 | 5.6 | 6.1 | 5.6 | 6.1 |
| 003 | 5.3 | 6.1 | 9.5 | 4.8 | 4.7 | 4.9 | 4.9 |
| 008 | 7.3 | 4.9 | 7.5 | 5.2 | 6.9 | 6.7 | 6.8 |
| 012 | 9.0 | 6.1 | 9.3 | 9.2 | 9.2 | 9.2 | 9.4 |
| 019 | 35.0 | 11.0 | 16.8 | 28.5 | 27.0 | 30.8 | 27.3 |
| 021 | 5.3 | 5.9 | 9.0 | 4.4 | 4.8 | 4.4 | 5.3 |
| 022 | 3.9 | 5.6 | 8.6 | 3.6 | 4.8 | 4.1 | 4.6 |
| 031 | 21.0 | 11.2 | 17.2 | 17.6 | 16.3 | 17.2 | 16.7 |
| 031 | 26.0 | 16.3 | 24.9 | 19.0 | 22.5 | 17.3 | 22.2 |
| 033 | 13.6 | 10.2 | 15.5 | 14.3 | 15.8 | 14.3 | 16.2 |
| Mean | 13.2 | 12.6 | 8.3 | 11.2 | 11.8 | 11.5 | 11.9 |
| SD | 10.6 | 5.7 | 3.7 | 8.4 | 8.1 | 8.2 | 8.1 |

ACH<sub>VENT</sub>: mechanical ventilation rate; ACH<sub>SS30</sub>: ventilation estimate by steady state CO<sub>2</sub> level with CO<sub>2</sub> generation at 0.3L/min per person; ACH<sub>SS46</sub>: ventilation estimate by steady state CO<sub>2</sub> level with CO<sub>2</sub> generation at 0.46L/min per person; ACH<sub>DI</sub>: ventilation rate estimate by CO<sub>2</sub> decay constants using dry ice; ACH<sub>BV</sub>: ventilation rate estimate by CO<sub>2</sub> decay constants using baking soda and vinegar; ACH<sub>DI63</sub>: ventilation rate estimate by time needed to remove 63% excess CO<sub>2</sub> by dry ice; ACH<sub>BV63</sub>: ventilation rate estimate by time needed to remove 63% excess CO<sub>2</sub> by baking soda and vinegar

**Table 4:** Ventilation rate estimates by time needed to remove 63% excess CO_2_ released by dry ice or baking soda and vinegar.

| RM# | Method | C<sub>R</sub><br>ppm | C<sub>S</sub><br>ppm | C<sub>E</sub><br>ppm | C<sub>63%E</sub><br>ppm | t<sub>2</sub><br>min | ACH<sub>T63%</sub><br>60/t<sub>2</sub> |
| --- | --- | --- | --- | --- | --- | --- | --- |
| 002 | DI | 403 | 3956 | 3553 | 1718 | 10.7 | 5.6 |
|  | BV | 410 | 2800 | 2390 | 1294 | 9.9 | 6.1 |
| 003 | DI | 416 | 3552 | 3136 | 1576 | 12.3 | 4.9 |
|  | BV | 412 | 1901 | 1489 | 963 | 12.3 | 4.9 |
| 008 | DI | 434 | 2459 | 2025 | 1183 | 9.0 | 6.7 |
|  | BV | 399 | 2290 | 1891 | 1099 | 8.8 | 6.8 |
| 012 | DI | 427 | 3064 | 2637 | 1403 | 6.5 | 9.2 |
|  | BV | 410 | 2340 | 1930 | 1124 | 6.4 | 9.4 |
| 019 | DI | 434 | 2901 | 2467 | 1347 | 2.0 | 30.8 |
|  | BV | 399 | 1112 | 713 | 663 | 2.2 | 27.3 |
| 021 | DI | 434 | 4265 | 3831 | 1852 | 13.5 | 4.4 |
|  | BV | 410 | 1782 | 1372 | 917 | 10.6 | 5.7 |
| 022 | DI | 427 | 4103 | 3676 | 1787 | 14.7 | 4.1 |
|  | BV | 399 | 2474 | 2075 | 1167 | 13.0 | 4.6 |
| 031 | DI | 427 | 2066 | 1639 | 1033 | 3.5 | 17.2 |
|  | BV | 416 | 2006 | 1590 | 1004 | 3.6 | 16.7 |
| 032 | DI | 427 | 2703 | 2276 | 1269 | 3.5 | 17.3 |
|  | BV | 410 | 2139 | 1729 | 1049 | 2.7 | 22.2 |
| 033 | DI | 434 | 2530 | 2096 | 1210 | 4.2 | 14.3 |
|  | BV | 432 | 1291 | 859 | 750 | 3.7 | 16.2 |

DI: dry ice. BV: baking soda and vinegar. C<sub>R</sub>: outdoor CO<sub>2</sub> level. C<sub>S</sub>: peak CO<sub>2</sub> level after CO<sub>2</sub> generation by dry ice or baking soda. C<sub>E</sub>: excess CO<sub>2</sub> generated by dry ice or baking soda (C<sub>S</sub> - C<sub>R</sub>). C<sub>63%E</sub>: CO<sub>2</sub> level after 63% excess CO<sub>2</sub> is removed (C<sub>S</sub>-63%C<sub>E</sub>). t<sub>2</sub>: time (min) needed to reach C<sub>63%E</sub>. ACH<sub>T63%</sub>: ventilation rate in air change per air based on t<sub>2</sub>

As ventilation rate is likely below 6 ACH in most dental treatment rooms in private dental practices in small free-standing buildings in the US[13], CO_2_ generation rate (G_P_) of 0.30 L/min is more appropriate for ventilation rate estimate using equation #1. We listed the corresponding steady state CO_2_ levels and ventilation rates in rooms with various sizes with 8-ft ceiling height using n = 3, G_P_ = 0.30 L/min, and C_R_= 400 ppm (Supplemental Table 1). DCPs may use the steady state CO_2_ levels measured in their dental treatment rooms to roughly estimate the ventilation rate using this table.

### Ventilation rates by CO_2_ decays using dry ice or baking soda

Results of ventilation estimates by CO_2_ decay using dry ice or baking soda and vinegar are shown in Table 3. Both methods appeared to be reliable in assessing the ventilation rates of dental treatment rooms. The CO_2_ decay curves demonstrate that CO_2_ levels decreased faster over time in rooms with high ACH_VENT_ (Figure 3 A, B). ACH_DI_ values ranged from 3.6 to 28.5 (11.2±8.4) and were highly correlated with the mechanical ventilation rates ACH_VENT_ (r=0.99, p<0.0001) (Figure 3 C). ACH_DI_ was lower than ACH_VENT_ (mean difference = 2.0, paired t=2.32, p=0.046). Similarly, ACH_BV_ ranged from 4.7 to 27.0 (11.8±8.1) and also correlated highly with ACH_VENT_ (r=0.98, p<0.0001) (Figure 3 D). There was no statistically significant difference between ACH_BV_ and ACH_VENT_ (mean difference = 1.4, paired t=1.48, p=0.174), or between ACH_BV_ and ACH_DI_ (mean difference = 0.59, paired t=1.26, p=0.239).

**Figure 3.**
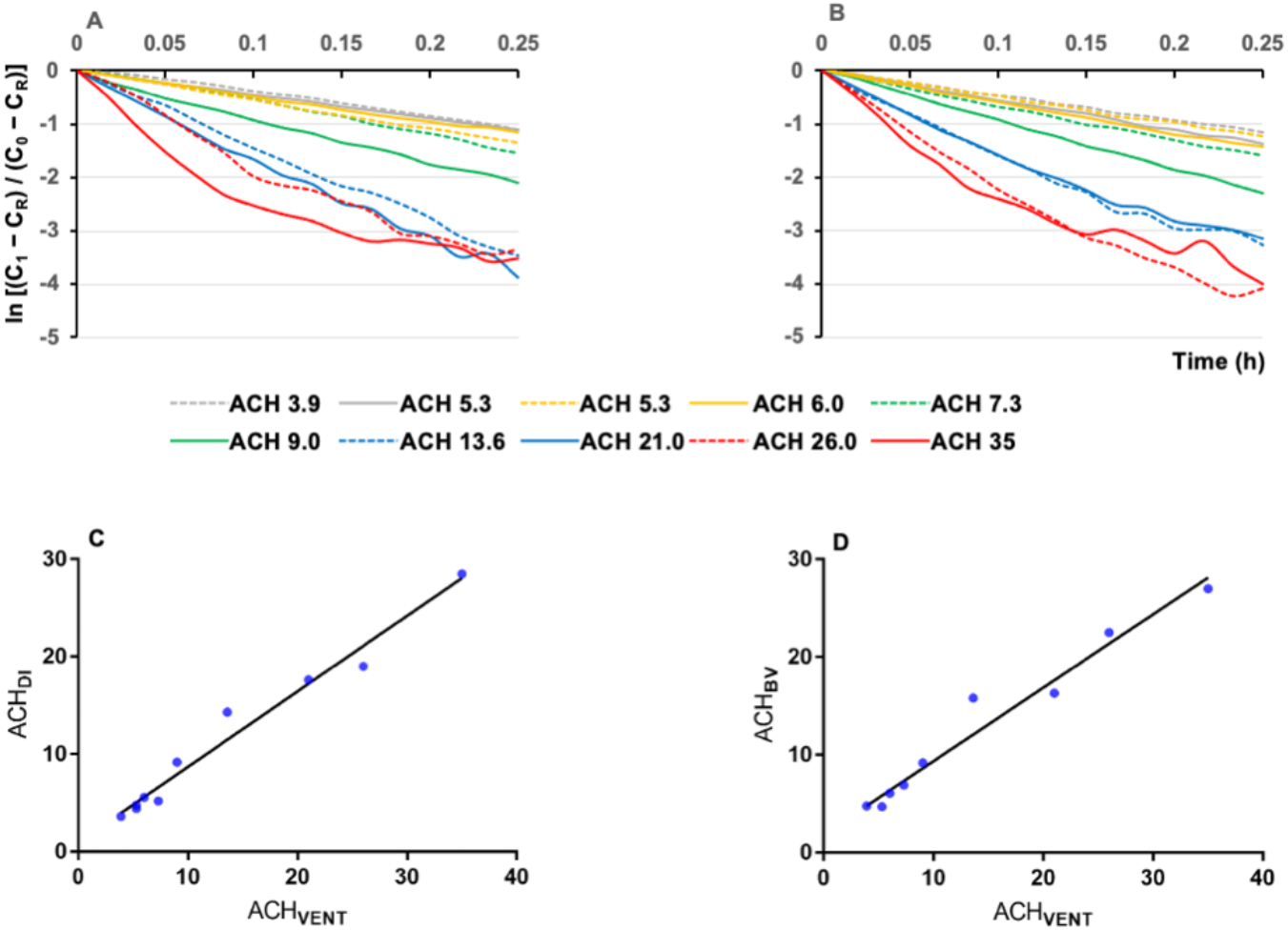
A: CO_2_ decay constants by dry ice, B: CO_2_ decay constants by baking soda and vinegar; C and D: association between mechanical ventilation rates measure by air flow (ACH_VENT_) and ventilation rates measured by CO_2_ decay using dry ice (ACH_DI_) and baking soda and vinegar (ACH_BV_) in dental treatment rooms. Rooms with high mechanical ventilation rates showed rapid decrease of CO_2_ concentrations over time (A, B). Both ACH_DI_ and ACH_BV_ are linearly correlated with ACH_VENT_ (C, D).

### Ventilation rates by time to 63% removal of excess CO_2_ generated by dry ice or baking soda

Ventilation rates calculated by time needed to remove 63% of excess CO_2_ generated by dry ice of baking soda are presented in Table 3. ACH_DI63_ values ranged from 4.1 to 30.8 (11.5±8.6) and were highly correlated with the mechanical ventilation rates ACH_VENT_ (r=0.98, p<0.0001) (Figure 4 A). There was no statistically significant difference between ACH_DI63_ and ACH_VENT_ (mean difference = 1.8, paired t=1.93, p=0.086).

**Figure 4.**
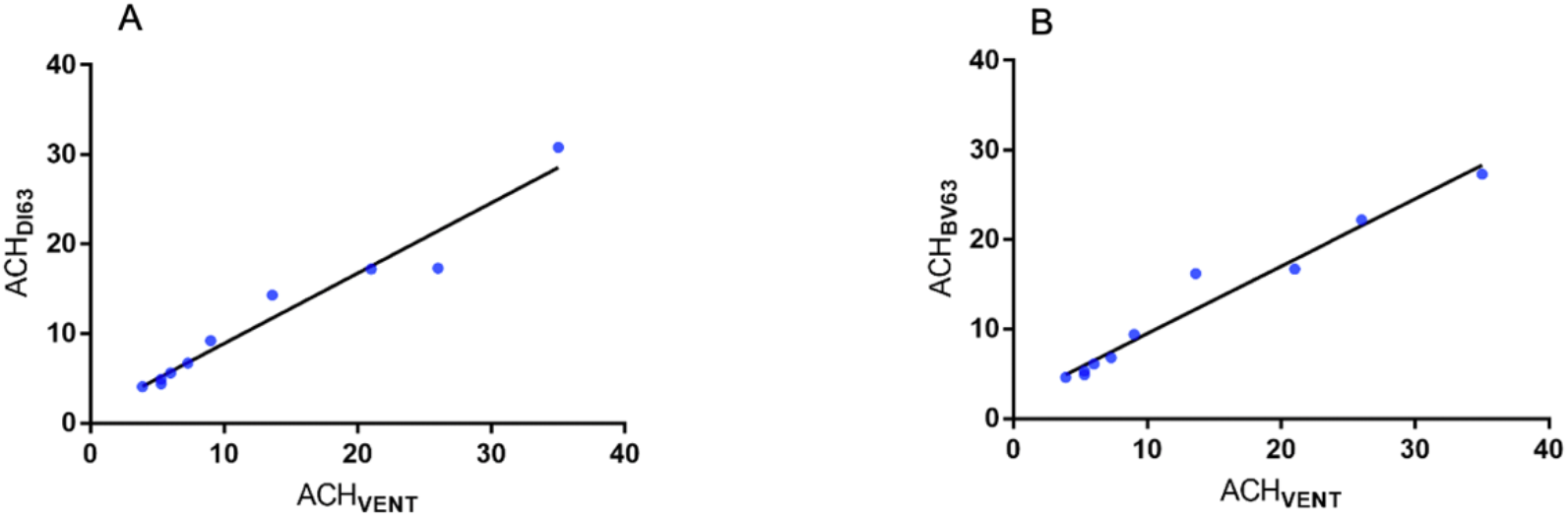
A: Correlations between ventilation rate by air flow (ACH_VENT_) and ventilation rates by time needed to reach 63% of excess CO_2_ generated using A: dry ice (ACH_DI63_) and B: baking soda and vinegar (ACH_BV63_).

Similarly, ACH_BV63_ ranged from 4.6 to 27.3 (11.9±8.1) and correlated highly with ACH_VENT_ (r=0.98, p<0.0001) (Figure 4 B). There was no statistically significant difference between ACH_BV63_ and ACH_VENT_ (mean difference = 1.3, paired t=1.34, p=0.213), or between ACH_BV63_ and ACH_DI63_ (mean difference = 0.50, paired t=0.76, p=0.467).

A Microsoft Excel template is provided in Supplemental Table 2 that will allow DCPs to calculate the ventilation rate of their offices by inputting the values of peak CO_2_ level (C_S_), outdoor CO_2_ level (C_R_) and time (min) needed to reach 63% removal of excess CO_2_ generated by dry ice or baking soda.

## Discussion

The findings of the present study indicate that dental operatories with low mechanical ventilation rates and overcrowding facilitate CO_2_ accumulations. Ventilation rates could be measured by assessing natural or experimental build-up of CO_2_ levels in dental treatment rooms using a consumer-grade CO_2_ sensor. We found that ventilation rates in ACH could be accurately assessed by observing the changes in CO_2_ levels after a simple mixing of household baking soda and vinegar in dental settings. Time needed to remove 63% of excess CO_2_ generated by baking soda could be used to accurately calculate the ventilation rates with the help of a basic calculator.

Our findings show that CO_2_ level may consistently stay above 800 ppm in rooms with ventilation rates below 6 ACH, especially when three or more persons (including the patient who was not wearing a mask) are in the room during dental treatments. We observed that CO_2_ level stayed above 1000 ppm and approached 1600 ppm when 3 to 6 persons were in a room with 3.9 ACH in clinical teaching scenarios involving dental implant treatment. High levels of CO_2_ indicate high concentrations of respiratory aerosols in the room. It is possible that these aerosols contain pathogens if the patient is not wearing a mask and is infected but asymptomatic or pre-symptomatic. Effective mitigation measures will be required in these rooms to improve the air quality even without the ongoing infectious disease pandemic. Overcrowding should be avoided in rooms with poor ventilation. In dental operatories with ventilation rates higher than 10 ACH, the CO_2_ levels stayed consistently below 700 ppm in most cases with 3 persons in the room. Our data demonstrated a clear dependency of CO_2_ levels on number of persons in the room and the mechanical ventilation rate. CO_2_ level is a proxy for indoor air quality as it represents the fraction of rebreathed air, or the proportion of inhaled air that was exhaled by others in the same indoor environment. Though numerous epidemiological studies indicate CO_2_ begins to have negative health effects at 700 ppm and respiratory symptoms may occur when indoor CO_2_ concentration is above 1000 ppm [25], our main concern is the concurrent accumulation of respiratory aerosols that may contain infectious disease pathogens. Numerous studies have shown that exhaled air from infected patients contains respiratory disease pathogens, including rhinovirus, influenza virus and Mycobacterium tuberculosis [4, 26-28]. Patients with early stages of COVID-19 may release millions of SARS CoV-2 viral copies per hour in exhaled air [29]. In a recent study that modeled factors associated with the spread of respiratory infectious disease in dental offices, CO_2_ levels were found to play the most important role on the risk of infectious disease transmission [11]. CO_2_ levels at 774 ppm was considered as low risk but those at or above 1135 ppm may increase the risk of disease transmission in dental offices [11].

It is important to point out that the setting for the current study is a postdoctoral dental training institution affiliated with an academic medical center, which may differ significantly in ventilation conditions from private dental practices that have a solo or a few dental practitioners. Ventilation conditions in different dental settings are largely unknown as the ventilation design of dental offices is not regulated as other outpatient healthcare facilities that are required to have 6 to 15 ACH by ASHRAE and CDC [30, 31]. Godwin and colleagues reported that ventilation rate was 1.12 ACH in dental operatories of a small dental clinic (2400 ft^2^) [13], which is significantly lower than the mean of 13 ACH in the present study but resembles more closely to the mean of 1.09 ACH in typical residential households in the US [32].

Accurate and reliable measurements of ventilation rate in various dental settings are important for risk assessment and for risk mitigation planning in an era of frequent infectious disease pandemics. Mechanical ventilation rate is assessed by quantifying the amount of outdoor air flowing into and out of an indoor space using highly sophisticated instruments operated by trained professionals [16]. Technical barriers may have contributed to the scarcity of information regarding ventilation in dental settings. Besides direct air flow measurements, ventilation rate could be estimated using CO_2_ as a tracer gas. CO_2_ in an indoor space could be built up to a significantly higher level than in outdoor air, either through natural generation by the occupants or through experimental release of the gas in the indoor spaces [5, 33, 34]. Analysis of the steady state CO_2_ levels or the rate of CO_2_ concentration decays, which is directly dependent on the outdoor air flow rate from the ventilation system, will allow an estimate of the ventilation rate of the indoor space. We found that modeling the steady state CO_2_ levels using equation #1 correlated reasonably well with the mechanical ventilation rate, but may either under- or overestimate the ventilation rate based on different assumptions of human CO_2_ generation rates. In comparison, the CO_2_ concentration decay method relied on actual CO_2_ levels measured at the beginning and the end of a decay period (equation #2) and provided more accurate assessments and better approximation to the mechanical ventilation rates. CO_2_ concentrations in dental operatories could be built up to a level of about 1500 to 2500 ppm in 2 minutes using either dry ice or baking soda and vinegar. CO_2_ decays could then be monitored using a CO_2_ sensor that logs data in one-minute intervals. There are many affordable consumer-grade CO_2_ sensors readily available and suitable for the purpose of observing CO_2_ level changes over a period of time, from several minutes to several hours depending on the mechanical ventilation rates. The CO_2_ sensor used in the present study was purchased online for $159 and appeared to be a reliable tool for monitoring indoor CO_2_ levels in dental settings.

Although ventilation rate in ACH could be calculated by fitting a linear regression line over time into the natural log scale of time-varying concentrations of CO_2_ levels (equation #2), we found that a simplified method (equation #4) provided equally if not more accurate estimate of ventilation rate. As it is known that one complete air change will replace 63% of the airborne contaminants in the room with outdoor air [22-24], ventilation rate could be easily calculated using the time needed to remove 63% of excess CO_2_ as a contaminant. For example, assuming CO_2_ level in outdoor air is 400 ppm, and peak CO_2_ level is 1500 ppm after placing dry ice or baking soda inside the dental office for 2 minutes, excess CO_2_ inside the dental office will be 1500 – 400 = 1100 ppm at peak. The CO_2_ level that represents 63% removal of excess CO_2_ is therefore 1500 – 63% ⨯1100 = 807 ppm. If it takes 15 min for the CO_2_ level to reach 807 ppm from the peak of 1500 ppm, ventilation rate in ACH will be 60/15 = 4; and if it takes 2 hours for the CO_2_ level to reach 807 ppm, ACH will be 60/120 = 0.5. This method will allow dental practitioners to accurately estimate the ventilation rate in ACH using a simple calculator.

Ventilation rates measured by the CO_2_ decay methods were approximately 15% lower on average than those measured by air flow sensors at the air supply inlets or exhaust returns of the ventilation system. This discrepancy is expected as indoor CO_2_ could only be removed by fresh outdoor air brought in by the ventilation system or by outdoor air infiltrated through leaky doors and windows [35]. As the mechanically ventilated air was not composed of 100%, but 60% outdoor air at the time of this study, the CO_2_ removal rate was understandably lower than the total air flow rate. In practical sense, ventilation rates measured by CO_2_ decay methods are better indicators of outdoor air flow, which is more important than the recycled indoor air flow measured by airflow sensors in terms of air contaminant removal efficiencies.

Our data showed that household baking soda (NaHCO_3_) and vinegar (5% acetic acid) could be used to generate CO_2_ in dental office to assess the ventilation rate by observing the CO_2_ concentration decays using a CO_2_ sensor and a basic calculator. This method will allow dental practitioners to reliably estimate the ventilation rate in their dental offices without expensive equipment and advanced technical skills. The test could be completed within 30 minutes in spaces with ventilation rate higher than 2 ACH but may take longer time if the ventilation is significantly below 1 ACH. We recommend to plan a two-hour observation time during off-hours with the building ventilation system operating in its normal setting.

We consider that it is very important for every dental practitioner to be able to accurately assess the ventilation rate in their working environments. Epidemiological data showed that transmission of COVID-19 is almost exclusively an indoor phenomenon, with 99.97% of the transmissions occurring in an indoor environment [29]. Airborne transmission through respiratory aerosols is increasingly recognized as a major driver for the COVID-19 pandemic [36-38]. As essential healthcare providers, dental professionals work in the frontline during the pandemic and need to adopt measures to mitigate the risk of aerosol transmission in addition to droplet and contact precautions that have been the standard of infection control in dental offices [39].

We recommend that mitigation measures be taken for dental treatment rooms that have a ventilation rate below 15 ACH, which is required for procedure rooms in outpatient healthcare facilities by CDC guidelines [30]. While in theory the most effective measure for air quality improvement in dental offices is to increase outdoor air flow rate through the mechanical ventilation system or through natural ventilation by opening doors and windows, such measure is severely limited by the weather or climate conditions and often impractical or impossible to realize in practice. An effective alternative is to improve air filtration using upgraded filters in the ventilation system and portable air cleaners (PAC) equipped with high efficiency particulate air (HEPA) filters. We showed that a PAC with a HEPA filter and a clean air delivery rate of 250 cubic feet per minute could add an equivalent of 17 ACH to an average sized dental operatory (10 ft L, 11ft W, 8 ft H) [17]. The PAC was especially effective in removing aerosols from rooms with low mechanical ventilation rate because of the lack of disturbance from the high air flow of the ventilation system [17]. For larger spaces where PAC is not practical or effective, upper room ultraviolet germicidal irradiation (UVGI) system may be considered as it has been shown to be effective in healthcare settings [40-42]. The upper room UVGI system requires professional installation and maintenance to ensure its safe operation.

In summary, we found that dental operatories with low ventilation rates (ACH_VENT_ ≤ 6) facilitate CO_2_ accumulations. Crowding inside the room contributed to elevated CO_2_ levels. Ventilation rates could be reliably calculated by observing the changes in CO_2_ levels after a simple mixing of household baking soda and vinegar in dental settings. Time needed to remove 63% of excess CO_2_ generated by baking soda could be used to accurately assess the ventilation rates using a consumer-grade CO_2_ sensor and a basic calculator. For rooms with ventilation rate below 15 ACH, we suggest the addition of a PAC with HEPA filter and proper clean air delivery rate to facilitate air quality improvement in dental treatment rooms.

## Data Availability

All data are available upon request

## Acknowledgements

The authors declare no conflict of interests. This study is supported in part by the Eastman Institute for Oral Health Foundation, Rochester, New York. We thank building engineers Dan Mateer and Kevin McLellan at Johnson Control Inc. and mechanical engineers Ray Richard and Karen Pembroke at Facility Operations, University of Rochester Medical Center for their technical expertise and assistance with the present study.

## Supplemental materials

### Using steady state CO_2_ level during dental treatments to estimate ventilation rates

The following table could be used to roughly estimate the ventilation rate of the dental treatment rooms in 3 steps. First, measure the length and width of the room to get the area in square feet (ft^2^). Second, determine the steady state CO_2_ level during a dental treatment procedure that lasts more than 10 minutes as follows: with the dentist, dental assistant and the patient together in the room and without any person entering or leaving the room, read the CO_2_ sensor readings 10 minutes into the procedure and record the next 5 readings at 1 min interval, add the 5 readings to get the sum and divide the sum by 5, the result is the steady state CO_2_ level. Third, match the steady state CO_2_ level to the closest number under the area column of your room size, the number in the ACH column in the same row is the ventilation rate estimate. For example, if the CO_2_ level reaches a steady state level of about 1062 ppm during a dental treatment that lasted longer than 5 minutes with 3 persons in a room that is 110 ft^2^ (10-ft W x11-ft L) in area, the ventilation rate is about 3 ACH. It will be about 6 ACH if the CO_2_ level stays at about 761 ppm (Supplemental Table 1).

**Supplemental Table 1:** Steady state CO_2_ levels and ventilation rate in air change per hour (ACH)*

| Area<br>(ft<sup>2</sup>) \ ACH | 100 | 110 | 120 | 130 | 140 | 150 | 160 | 170 | 180 | 190 | 200 |
| --- | --- | --- | --- | --- | --- | --- | --- | --- | --- | --- | --- |
| 1 | 2784 | 2567 | 2386 | 2234 | 2103 | 1989 | 1890 | 1802 | 1724 | 1655 | 1592 |
| 2 | 1592 | 1484 | 1393 | 1317 | 1251 | 1195 | 1145 | 1101 | 1062 | 1027 | 996 |
| 3 | 1195 | 1122 | 1062 | 1011 | 968 | 930 | 897 | 867 | 841 | 818 | 797 |
| 4 | 996 | 942 | 897 | 858 | 826 | 797 | 772 | 751 | 731 | 714 | 698 |
| 5 | 877 | 833 | 797 | 767 | 741 | 718 | 698 | 680 | 665 | 651 | 638 |
| 6 | 797 | 761 | 731 | 706 | 684 | 665 | 648 | 634 | 621 | 609 | 599 |
| 7 | 741 | 710 | 684 | 662 | 643 | 627 | 613 | 600 | 589 | 579 | 570 |
| 8 | 698 | 671 | 648 | 629 | 613 | 599 | 586 | 575 | 566 | 557 | 549 |
| 9 | 665 | 641 | 621 | 604 | 589 | 577 | 566 | 556 | 547 | 539 | 532 |
| 10 | 638 | 617 | 599 | 583 | 570 | 559 | 549 | 540 | 532 | 525 | 519 |
| 11 | 617 | 597 | 581 | 567 | 555 | 544 | 535 | 527 | 520 | 514 | 508 |
| 12 | 599 | 581 | 566 | 553 | 542 | 532 | 524 | 517 | 510 | 505 | 499 |
| 13 | 583 | 567 | 553 | 541 | 531 | 522 | 515 | 508 | 502 | 497 | 492 |
| 14 | 570 | 555 | 542 | 531 | 522 | 514 | 506 | 500 | 495 | 490 | 485 |
| 15 | 559 | 544 | 532 | 522 | 514 | 506 | 499 | 493 | 488 | 484 | 479 |

\*Based on ventilation rate $ACH = 6 \times 10^4 n G_P / [V(C_{SS}-C_R)]$ , where number of persons in the room $n=3$ , CO<sub>2</sub> generation rate per person $G_P=0.3L/min$ , outdoor CO<sub>2</sub> level $C_R=400$ ppm, and $V$ is the volumetric size of the room with ceiling height = 8-ft.

### Calculating ventilation rate using CO_2_level change after mixing baking soda and vinegar

Supplemental Table 2 is a dynamic template that will allow you to enter 3 values to get the ventilation rate in air change per hour (ACH) for your treatment rooms: 1, the peak CO_2_ level (C_S_), 2, the outdoor CO_2_ level, and 3. Time needed to reach 63% removal of excess CO_2_.

**Supplemental Table 2:** Ventilation rate estimate using time needed to remove 63% excess CO_2_ generated by baking soda and vinegar.

|  |  |  |  |
| --- | --- | --- | --- |
| CO<sub>2</sub> at peak | 2800 | ppm | B1. C<sub>s</sub> = Peak CO<sub>2</sub> level C<sub>s</sub> |
| CO<sub>2</sub> outdoors | 400 | ppm | B2. C<sub>R</sub> = Outdoor CO<sub>2</sub> level C<sub>R</sub> |
| Excess CO<sub>2</sub> | 2400 | ppm | B3. C<sub>E</sub> = Excess CO<sub>2</sub>. C<sub>E</sub> = C<sub>s</sub> - C<sub>R</sub> |
| After removal of 63% excess CO<sub>2</sub> | 1288 | ppm | B4. C<sub>63%</sub> = CO<sub>2</sub> level after removal |
| Time needed to remove 63% excess CO<sub>2</sub> | 15.0 | min | B5. Time to reach C<sub>63%</sub> (Value in min) |
| Ventilation rate | 4.0 | h<sup>-1</sup> | B6. Air change per hour, or ACH |

To calculate air change per hour, 3 values need to be entered into the above template:

## Notes

### Competing Interest Statement

The authors have declared no competing interest.

### Funding Statement

This study was supported in part by the Eastman Institute for Oral Health Foundation, Rochester, New York, USA

### Author Declarations

University of Rochester Medical Center Research Subject Review Board

